# Sudden hyposmia as a prevalent symptom of COVID-19 infection

**DOI:** 10.1101/2020.04.06.20045393

**Authors:** Rosario Marchese-Ragona, Giancarlo Ottaviano, Nicolai Piero, Andrea Vianello, Carecchio Miryam

## Abstract

Severe Acute Respiratory Syndrome Coronavirus-2 (SARS-CoV-2) has recently caused a pandemic that has involved Italy as the second worldwide nation in terms of infected patients and deaths. The clinical manifestation of Covid-19 ranges from asymptomatic carrier status to severe pneumonia. Asymptomatic individuals in Covid-19 are those who are carriers of the virus but do not show clinical symptoms and are able to transmit the disease in the same degree as symptomatic carriers. In order to contain contagions is of supreme importance to identify asymptomatic patients because this subpopulation is one of the main factors contributing to the spread of this disease. We report on six Italian patients with COVID-19 who presented sudden hyposmia as the only or most prominent disease manifestation, without upper or lower respiratory tract involvement or other major features of the disease. A supra-threshold olfaction test confirmed the hyposmia in all patients. The onset of hyposmia during a Covid-19 outbreak should be considered as a warning sign of an infection that requires a diagnostic test for Covid-19

---

Since December 2019, a novel coronavirus SARS-CoV-2 (Covid-19) outbreak emerged in Wuhan, China, and subsequently rapidly spread to several countries. As of March 26, 2020, 533.416 cases with 24.082 deaths have been confirmed worldwide. Currently, United States, China, and Italy respectively are the countries with the highest number of cases^1^. In a study of 7,736 Covid-19 patients in China, of all the clinical symptoms, hyposmia was not reported in any patient^2^. We report on 6 subjects from Padua, Italy, who presented with hyposmia as the only or main manifestation of Covid-19. All patients were considered as “healthy people” before they were screened because of close contact with a lab-confirmed case of Covid-19. None of the patients reported acute, chronic or seasonal upper airway disease before infection, in one case a four-day fever was present after the onset of hyposmia; two patients reported myalgia the day before the onset of hyposmia and a mild dry cough after the hyposmia. Since olfactory disorders have a significant impact on taste, hypogeusia was reported in almost all cases^3^. This study was approved by the local ethics committee and all subjects gave informed consent to participate in the study. Patient data and clinical features are summarized in Table 1. All patients underwent “le nez du vin” a supra-threshold six odours smell test that confirmed hyposmia in all cases^4^. The clinical manifestation of Covid-19 ranges from asymptomatic carrier status to severe pneumonia. Asymptomatic individuals in Covid-19 are those who are carriers of the virus but do not show clinical symptoms. Zou et al. documented that the viral load found in asymptomatic patients was similar to that found in symptomatic patients. Asymptomatic carriers of COVID-19 virus are able to transmit the disease in the same degree as symptomatic carriers^5^. In order to contain contagions is of supreme importance to identify asymptomatic patients because this subpopulation is one of the main factors contributing to the spread of this disease.

**Table 1.** Patient data and clinical features.

| Patient no | Age | Sex | Fever | Cough | Cold | Sore throat | Myalgias | Hypo-smia | Hypo-geusia | Other diseases | Smoke | Upper airways diseases | Smell test (wa) | Tested for |
| --- | --- | --- | --- | --- | --- | --- | --- | --- | --- | --- | --- | --- | --- | --- |
| 1 | 37 | F | N | N | N | N | Y | Y | Y | N | N | N | 4/6 | SC |
| 2 | 50 | F | N | N | N | N | N | Y | Y | N | N | Y (allergic rhinitis) | 4/6 | SC |
| 3 | 25 | F | N | N | N | N | Y | Y | Y | Hypo-thyroidism | Y * | N | 2/6 | # |
| 4 | 24 | M | N | N | N | N | N | Y | Y | N | N | N | 4/6 | SC |
| 5 | 29 | M | N | N | N | N | N | Y | Y | N | N | N | 4/6 | SC |
| 6 | 29 | F | Y (max 39.6°C) | Y | N | N | N | Y | N | N | N | N | 4/6 | SC |

\*non-daily smoker (less than 5 cigarettes a month; wa=Nez du Vin wrong answer; SC= strict contact with lab-positive covid-19; # Patient number 3, complained only of persistent hyposmia without close contact with any Covid-19 case; she insisted on being tested because she had heard through social media a Covid-19 patient presenting only hyposmia (the patient number 1!).

The onset of hyposmia during a Covid-19 outbreak should be considered as a warning sign of an infection that requires a diagnostic test for Covid-19 and the detection and quarantine of patient close contacts. More extensive studies will be needed to systematically assess the frequency of hyposmia among Covid-19 patients, its pathogenesis, duration and potential role as a marker of disease progression or severity.

## Data Availability

All data generated or analysed during this study are included in this article

## Notes

### Competing Interest Statement

The authors have declared no competing interest.

### Funding Statement

no external funding was received

## References

1. Johns Hopkins Center for Systems Science and Engineering 2019-nCoV global cases. https://gisanddata.maps.arcgis.com/apps/opsdashboard/index.html#/bda7594740fd40299423467b48e9ecf6 Date accessed: March 27, 2020

2. Guan WJ, Ni ZY, Hu Y, et al. Clinical Characteristics of Coronavirus Disease 2019 in China. N Engl J Med 2020, Feb 28. doi:10.1056/NEJMoa2002032

3. Doty RL. The olfactory system and its disorders. Semin Neurol 2009;29:74–81

4. McMahon C, Scadding GK. Le Nez du Vin - a quick test of olfaction. Clin Otolaryngol Allied Sci 1996;21:278–80

5. Zou L, Ruan F, Huang M, et al. SARS-CoV-2 Viral Load in Upper Respiratory Specimens of Infected Patients. N Engl J Med. 2020 Mar 19;382(12):1177–1179.

